# Increasing Representativeness in the *All of Us* Cohort Using Inverse Probability Weighting

**DOI:** 10.1101/2024.10.02.24314774

**Authors:** Manoj S. Kambara, Shivam Sharma, John L. Spouge, Barry I. Graubard, I. King Jordan, Leonardo Mariño-Ramírez

## Abstract

Large-scale population biobanks rely on volunteer participants, which may introduce biases that compromise the external validity of epidemiological studies. We characterized the volunteer participant bias for the *All of Us* Research Program cohort and developed a set of inverse probability (IP) weights that can be used to mitigate this bias. The *All of Us* cohort is older, more female, more likely to have higher education, more likely to be covered by health insurance, less White, less likely to drink or smoke, and less likely to report being healthy compared to the US population. IP weights developed via comparison of a nationally representative database reduced the observed biases for all demographic and lifestyle characteristics. Furthermore, IP weighting corrected for differences in the correlation structure of the data. For all variables we corrected for, IP weighting brought correlation coefficients and pairwise variable associations closer to the nationally representative estimates. We provide our IP weights as a community resource to increase the representativeness and external validity of the *All of Us* cohort.

## Introduction

An important aim of epidemiological studies is to study the risk and occurrence of disease in whole populations, and to do this, studies utilize samples that are intended to be representative of a target population (TP), e.g., the U.S. population. Randomly sampling individuals from the TP with proper weighting (for the sampling rates with other adjustments) of the data from those who agree to participate ensures that results of analyses are generalizable or sometimes referred to representative of the TP, i.e., approximately unbiased for estimation and inferences of parameters for the target population. The *All of Us* Research Program is a study that aims to recruit a large and diverse cohort of the US population^1, 2^ from which data on demographics, social determinants of health, genetic factors, and health outcomes are collected.^2-4^ Unfortunately, the participants that make up the *All of Us* cohort are volunteers, i.e., they are not randomly selected from the TP and thus may differ from the TP with respect to demographic characteristics, disease risk factors, and health outcomes.^5, 6^ This volunteer participation bias means that inferred associations between risk factors and health outcomes for the *All of Us* cohort may not be externally valid for the TP.^5-8^

This problem of biased analyses of volunteer samples has been documented in other large biobank studies,^7, 8^ most notably the UK Biobank (UKBB).^9^ The UKBB cohort was also recruited from volunteers that contributed data, and thus is not generalizable to the United Kingdom TP.^10^ To improve generalization of the UKBB cohort inverse probability weighting (IPW) was studied.^11, 12^ IPW is a method that involves weighting individuals by the inverse of their estimated participation probability. For example, the UKBB cohort participants were more likely to be older (among other traits) compared to the UK population.^10, 11^ Thus, older participants had a higher probability of participation comparatively to younger participants. Given this higher participation probability among the older participants, these participants are inversely weighted down relative to the younger participants to reduce their overrepresentation in analyses. IPWs are constructed on multiple traits/variables whose joint distributions are known or can be estimated for the TP from external data sources, e.g., a census or valid survey of the TP. Using the IPW method, the UKBB cohort sample has been weighted to better match the UK population on demographic characteristics.^11, 12^

The recruitment process of the *All of Us* cohort differs from that of the UKBB cohort. The UKBB issued mailers to the UK population to obtain volunteers for the cohort that contributed their health data without any specific health-related or demographic criteria apart from age and location.^9, 13^ In contrast, the *All of Us* program leveraged two means of recruitment, healthcare provider organizations (HPOs) and direct volunteers, with a focus on the recruitment of populations previously underrepresented in biomedical research.^14^ We hypothesized that the *All of Us* cohort may also show volunteer participant bias, albeit in a different manner observed for the UKBB. If the *All of Us* cohort also shows volunteer participant bias, then IPW could be used, incorporating appropriate covariates measured in both the cohort and known or estimated in the TP, to mitigate this bias. The first aim of this study was to evaluate the representativeness and potential participation bias in the *All of Us* cohort by quantifying differences between *All of Us* participants and the US population for a variety of demographic, social, lifestyle, and health-related characteristics. The second aim of this study was to develop IPW to improve the generalizability of the *All of Us* cohort to the U.S. population. The third aim of the study was to empirically show that when using the IPWs for the *All of Us* cohort data that the results were closer to results from representative U.S. data.

## Methods

### Data Sources

#### National Health and Nutrition Examination Survey (NHANES)

The National Health and Nutrition Examination Survey (NHANES) is designed and conducted by the National Center for Health Statistics (NCHS), the Centers for Disease Control and Prevention (CDC),^15, 16^ to assess the health and nutritional status of adults and children in the United States. The survey examines a nationally representative sample each year. NHANES uses a stratified multistage cluster sample design to randomly select individuals to participate. The NHANES 2017 – March 2020 oversampled Hispanics, non-Hispanic Blacks, non-Hispanic Asians,^15^ individuals at or below 185% of the poverty level, and individuals aged 0-11 or 80 and over.^15^ The NHANES August 2021–August 2023 did no oversampling by race, Hispanic origin, and income, but there was oversampling for individuals aged 0-19 or 60 and over.^19^ NHANES calculates their own sample weights to ensure U.S. national generalizability of weighted analyses.^15^

NHANES collects demographic, socioeconomic, dietary, and health-related data from participants in two ways, an in-home interview and few weeks later in a mobile examination center, in which health examination and biospecimen collection is conducted.^15^ We combined the NHANES 2017 – March 2020 and August 2021–August 2023 samples where the sample weights for the combined samples were adjusted according to NCHS recommendations.^17^ The time period of the collection of the NHANES and the *All of Us* data were closely contemporaneous. The NHANES 2017 – March 2020 sample had a final interview response rate of 51.0% and a final examination response rate of 46.9%. ^15^ The NHANES August 2021–August 2023 sample had a final interview response rate of 34.6% and a final examination response rate of 25.7% with these lower response rates due to the pandemic.^16^ The total NHANES sample size of participants aged 18-79 was n=16,639; see **Supplementary Figure 1** for a flow chart of the NHANES sample.

Since there were mostly small rates of missing data for individual variables except for alcohol consumption with 21.41% missing in the NHANES dataset, we used a single imputation by chain equations, i.e., MICE, to impute for missing data values; see **Supplementary Methods** for further details of the imputation and **Supplementary Table 1** and **Supplementary Figure 2** for results of the imputation.^18^

#### All of Us

The *All of Us* Research Program cohort is a large-scale biobank resource including demographic, social, and genetic factors for adults within the United States.^1-4^ The *All of Us* Research Program is comprised of volunteers from diverse backgrounds. This study was conducted using version 7 of the *All of Us* Controlled Tier Dataset. This dataset includes individuals from the initial *All of Us* recruitment period, May 2018, to a data collection cutoff date of July 1^st^, 2022. *All of Us* recruits adults who are aged 18 or older and excludes those who are incarcerated at the time of enrollment or do not have the capacity to consent. Changes to Recruitment during the COVID-19 shifted to accommodate less in-person interaction.^19^ Participants were asked to answer an additional survey, called the COVID-19 Participants Experience (COPE), regarding their experience during the COVID-19 pandemic.^20^

We initially included all participants with at least one instance of the 10 variables in the initial cohort (n=379,454). Following this, we subset the cohort such that there was no missing data, leaving a final cohort of (n=310,533); see **Supplementary Figure 3** for flow chart of the data used for analysis.

#### Variable Harmonization

We harmonized the coding of the ten common variables between the *All of Us* and NHANES participant datasets used for developing the IP. These variables included age in years, categorical variables for race and ethnicity, sex at birth, birthplace, highest grade completed, marital status, health insurance, smoking habits, drinking habits, and self-reported overall health. The original and harmonized questions and answers are given in **Supplementary Table 2**.

### Inverse Probability Weight Calculation

#### Inverse Probability Weight Calculation

We used a logistic LASSO regression implemented with glmnet software to predict the probability of *All of Us* participation in order to calculate the IPW for each participant based on the ten variables included in the logistic model.^21^ We included the sample weights from the NHANES in the estimation of the logistic regression. To predict the IPW, we included each possible two-way interaction terms among the dummy and continuous variables in the logistic regression modeling. LASSO performs variable selection to include the predictors that contribute the most to participation. We predicted participation probability (*P_i_*) for each individual participant within the *All of Us* cohort. Using this probability, we calculated the raw IPW for each individual participant using the following formula: 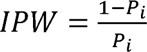.

#### Post-stratification

Following initial IPW calculation, these weights were adjusted by post-stratification using the postStratify function in the survey package in R.^22^ In order to post-stratify we utilized the same post-stratification cells that are used in NHANES. These post stratification cells were constructed from a complete cross classification of the American Community Survey estimated US population totals categories of age (18-39, 40-49, 50-59, 60-79), sex at birth (male or female), and race and ethnicity (White, Black or African American, Hispanic or Latino, Asian, Other/Multiracial) for 2017 - March 2020.^23^ For example, one cell used was the population count for White, male, and age 18-39. Post-stratification cells can be found in **Supplementary Table 3**. These post-stratified IPW, called IPWps, were used in the analyses.

### IPWps Validation

#### Population mean and proportion recovery

We initially validated the performance of the IPWps by measuring the recovery of the nationally representative NHANES population means and proportions for the ten variables, with the weights applied to the *All of Us* cohort. Using the following weighted mean estimator, 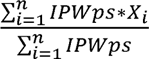, where *X_i_* is the variable of interest, we estimated means and proportions for the *All of Us* cohort to compare to that of NHANES.

#### Correlation coefficients

We measured the differences in correlation between all the ten variables in the NHANES cohort, *All of Us* cohort (both unweighted and weighted). The degree to which the *All of Us* weighted correlation coefficients moved closer to the NHANES correlation coefficient was used as an indication of reduction in participation bias. We measured the correlation between numeric and categorical variables using point biserial correlation, and the correlation between categorical variables using Cramer’s V.^24^

#### Pairwise variable associations

To further measure examine reduction in participation bias, we measured the differences in pairwise associations between all ten variables in the NHANES cohort, compared to pairwise associations in both the unweighted and weighted *All of Us* cohorts. We did this by running one-to-one logistic regressions for each pairwise combination of variables used in the IPWps weighting. To do this we converted each categorical variable to dummy variables, and then doing logistic regression for each combination. For example, we measured the correlation between one level of the “Overall Health” variable - “Overall Health (Very good)” – and one level of the “Drinking Habits” variable - “Drinking Habits (Twice or more a week)”. We then roughly calculated the confidence intervals for the pairwise variable associations using the confint function in R.

## Results

### Sample Makeup

For a nationally representative sample to compare the *All of Us* cohort against, we used data from the 2017 – March 2020 and the August 2021 - 2023 National Health and Nutrition Examination Survey (NHANES) run by the Centers for Disease Control (CDC).^15, 16^ We started with a total NHANES sample of n=27,493 participants, with data for ten demographic, social, and lifestyle characteristics common with *All of Us* participant data (**Supplementary Figure 1**). After restricting the age range and imputing missing data, we ended with a final sample of n=16,639 participants. For the *All of Us* dataset, we started with an initial cohort of n=379,454 participants, with data for at least one of the ten demographic, social, and lifestyle variables (**Supplementary Figure 3**). After exclusion of *All of Us* participants with missing data among the ten variables, and restricting the age range, we ended with a final sample of n=310,533. Details on the study cohort construction, and the harmonization of variables between the NHANES and *All of Us* cohorts, can be found in the Methods section and **Supplementary Table 2**. Details on imputation of NHANES data can be found in the Methods section and results of the imputation can be found in **Supplementary Table 1**.

### *All of Us* is different from the US population

We found that the *All of Us* participant cohort differed substantially from the US population. Compared to participants from the nationally representative NHANES cohort, *All of Us* participants differed for all ten characteristics considered here. *All of Us* participants are older, more female, more likely to have higher education, more US born, less married, more likely to be covered by health insurance, smoke less frequently, drink less alcohol, and self-report a lower overall general health than the US population. The differences between the nationally representative NHANES sample and the *All of Us* cohort can be seen in **Table 1**. Nearly all variables show significant differences between NHANES and *All of Us*.

**Table 1.**
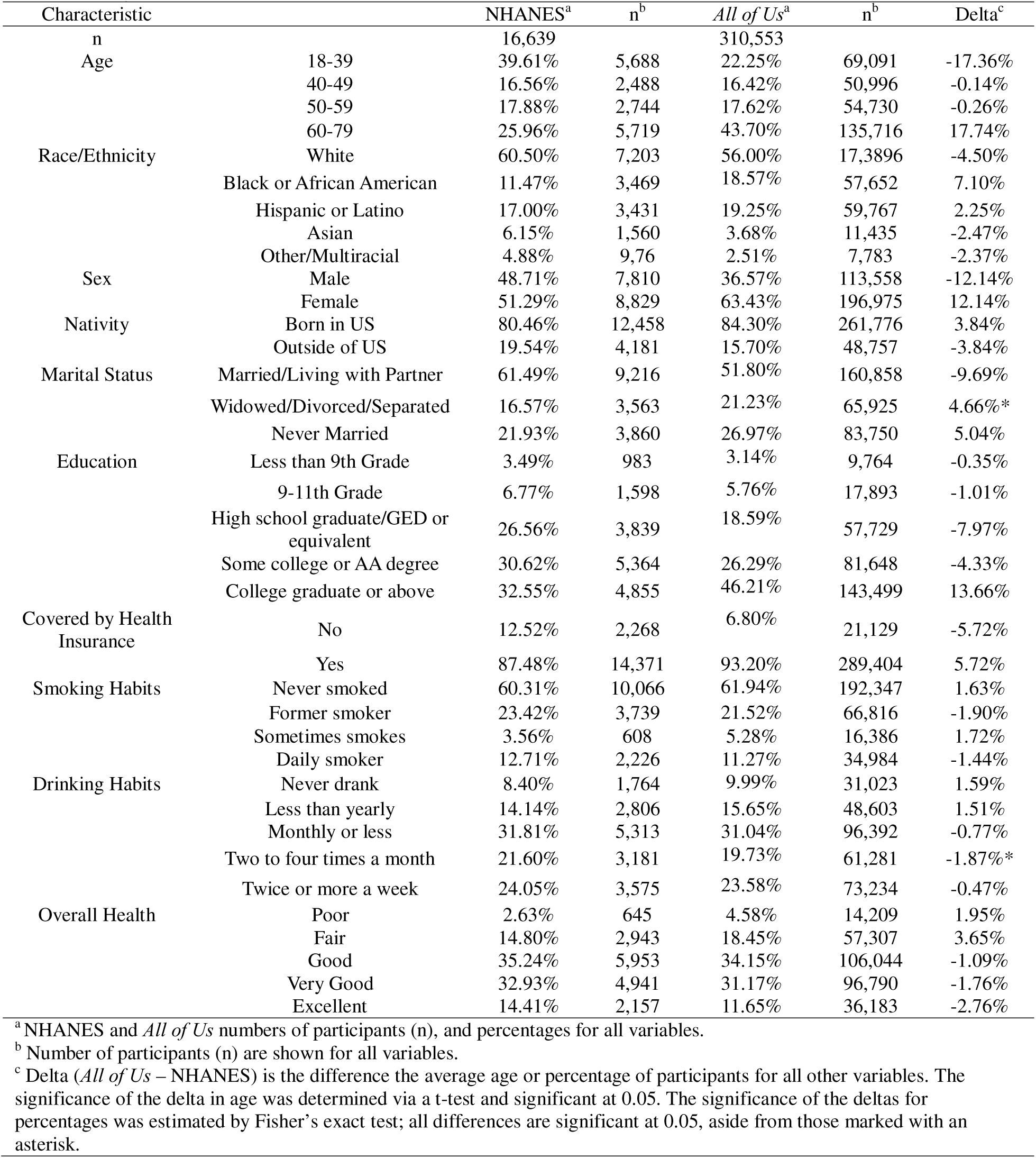
*All of Us* and NHANES participant comparison.

### Inverse probability (IP) weights for *All of Us*

To calculate IPW for *All of Us* participants, we developed a LASSO regression model that predicts the *All of Us* participation probability based on the ten harmonized variables for the *All of Us* cohort and the NHANES cohort. The IPW model also included all possible two-way interactions between the ten variables. Using the participation probabilities predicted from this model, we derived IPW for *All of Us* participants. Participants with characteristics that are overrepresented in the *All of Us* cohort compared to the US population are assigned lower IPW, whereas participants with underrepresented characteristics are assigned higher IPW. The process of variable selection for the LASSO regression IPW model can be found in **Supplementary Figure 4**. Once we calculated these IPW, we post-stratified the weights to match the sex, race and ethnicity, and age of the US population based on population counts from the ACS 2017 – March 2020 data.^23^ These counts can be seen in **Supplementary Table 3**. These post-stratified IPW, called IPWps, were used in the analyses. This ensured that the proportions for sex, race and ethnicity, and age matched exactly that of the US population. Results for each stage of weight development can be found in **Supplementary Table 4**.

To evaluate the performance of IPWps, we first tested if the weights reduced the effect of the ten participant variables on cohort participation within a univariate regression model. **Figure 1A** shows that the effects of all variables were greatly reduced when IPWps are included in the model, showing that the weighting was effective. Next, we found that IPWps for the *All of Us* cohort effectively recovered US population means and proportions for all variables included in the model. **Figure 1B** shows the effect of IPWps on all 10 variables included in weight development, and the values for these variables can be found in **Supplementary Table 4**.

**Figure 1.**
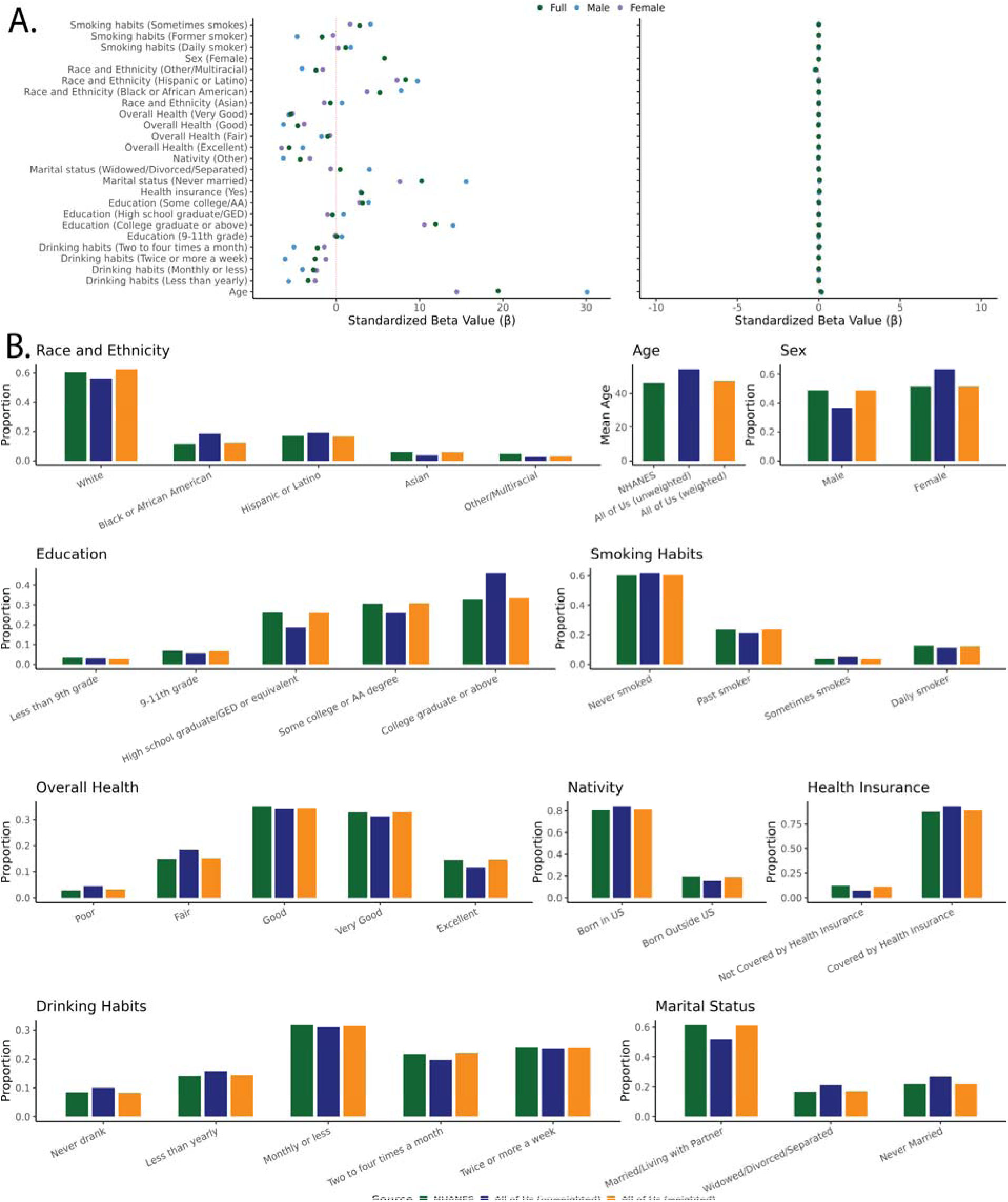
Inverse probability (IP) weights for the *All of Us* cohort. (A) Beta coefficients of variables prior to and after applying IPWps to univariate LASSO regression models. (B) Participant means and proportions for all 10 variables from weight development for the NHANES (green), *All of Us* unweighted (blue), and *All of Us* weighted (orange) cohorts.

### *All of Us* correlation structure

We evaluated the correlation structure of these variables by measuring the pairwise variable associations for a one-to-one logistic regression between each variable included in calculating the IPWps. **Figure 2A** shows the 30 correlations with the largest difference in pairwise variable associations between the unweighted *All of Us* cohort and the NHANES cohort. As seen in **Figure 2A**, the application of weights tended to bring pairwise variable associations closer to that of the nationally representative NHANES cohort. To evaluate if this effect was systemic for all combinations of variables, we regressed the pairwise variable associations for the unweighted and weighted *All of Us* cohorts against the nationally representative NHANES cohort (**Figure 2B and 2C**). While the unweighted *All of Us* cohort showed a fairly strong correlation with the NHANES cohort (R^2^ = 0.74), the weighted *All of Us* cohort showed a substantially stronger correlation (R^2^ = 0.96). All pairwise variable associations for all correlations included can be found in **Supplementary Table 5**. We also evaluated the correlation coefficients between all variables included in weighting and included these results in **Supplementary Figure 5** and **Supplementary Table 6**.

**Figure 2.**
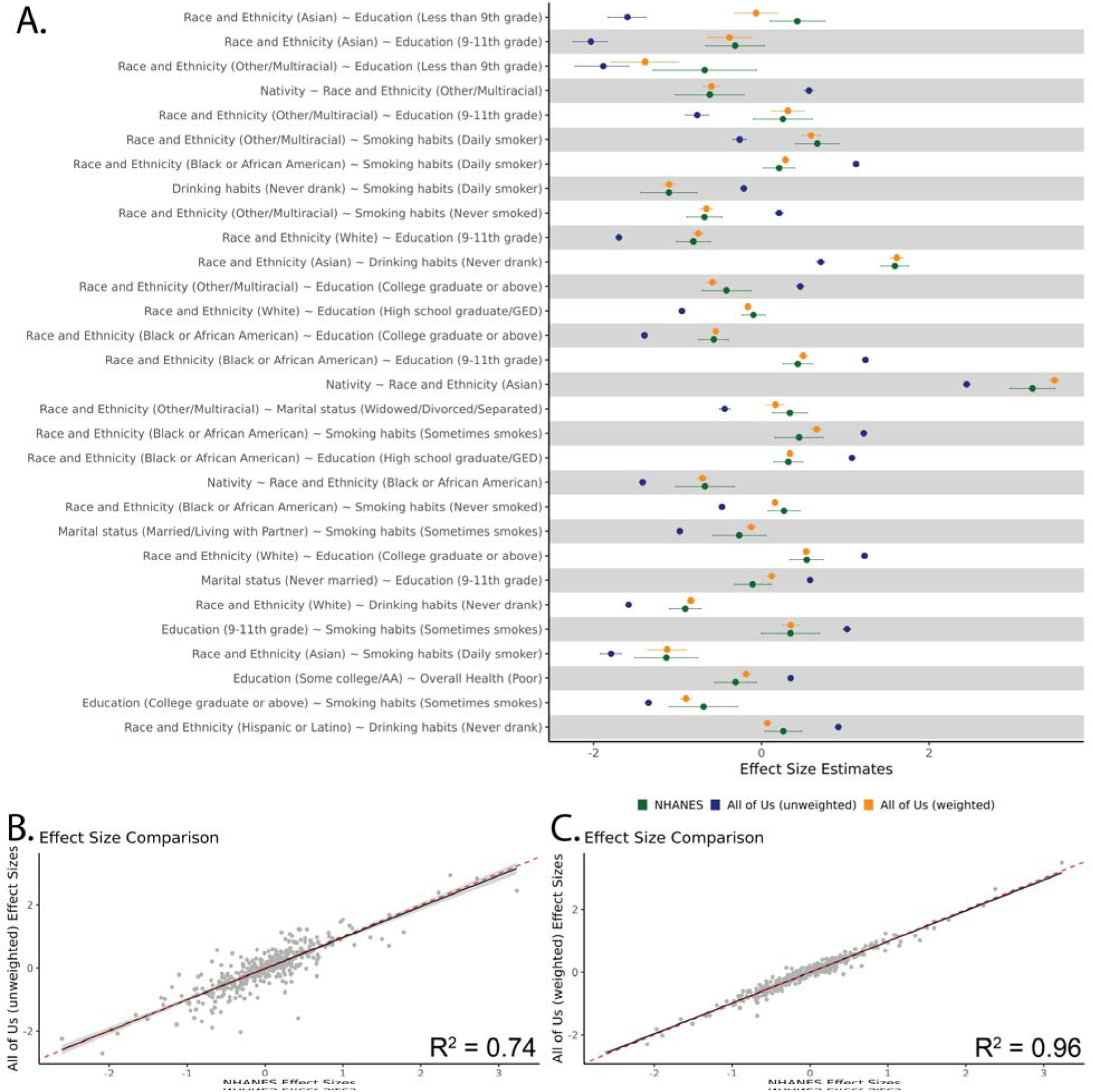
Pairwise regression of variables. (A) We ran one-to-one logistic regressions on each trait within the NHANES (green), *All of Us* unweighted (blue) and weighted (orange) cohorts. Here we plot the pairwise variable associations from the regression for the predictor variable with a 95% confidence interval. We show these for the 30 pairwise variable associations that showed the largest initial difference between the unweighted *All of Us* cohort and NHANES. We then regressed all pairwise variable associations for both the unweighted and weighted *All of Us* cohort against the NHANES cohort. (B) *All of Us* unweighted pairwise variable associations (y-axis) regressed against NHANES pairwise variable associations (x-axis). (C) *All of Us* weighted pairwise variable associations (y-axis) regressed against NHANES pairwise variable associations (x-axis). For B and C, the linear trend line (black) and ribbon (gray) are shown along with the y=x unity line (red, dashed).

## Discussion

The *All of Us* cohort has become a valuable resource for large-scale genetic epidemiology studies on diverse participant cohorts.^1, 3, 4^ Nevertheless, the extent to which the *All of Us* participant cohort reflects the demographic, social, lifestyle, and health outcome characteristics of the broader US population has yet to be systematically evaluated. In this study, we showed that the *All of* Us cohort differs substantially from the US population owing to volunteer participant bias. Similar to what has been seen for the UK Biobank, *All of* Us participants tend to be older, more educated, and more female than the US population.^5, 6, 10^ However, the *All of Us* cohort does not have the same healthy volunteer bias that has been observed for other biobank cohorts.^1^ On the contrary, the *All of Us* cohort shows higher prevalence values for a wide variety of disease categories compared to the US population.^1^ Taken together, these findings suggest that the results of disease association studies conducted on the *All of Us* cohort may not be externally valid for the US population.

In light of this problem, we developed a set of inverse probability weights (IPWps) to increase the population representativeness of the *All of Us* cohort, thereby supporting the external validity of epidemiological studies conducted using the database. Application of these IPWps to the *All of Us* cohort showed a reduction in observed participation bias and brought the demographic, social, and lifestyle characteristics in line with the US population. Furthermore, the IPWps we developed appeared to improve correlation structure among variables seen for the US population.

While the IPWps we developed are encouraging for improving US population generalization of the *All of Us* cohort, there are important limitations to consider. The IPWps were limited to matching based on only ten participant characteristics that were common to, and could be harmonized between, the nationally representative NHANES cohort and the *All of Us* cohort. There may be other variables that affect participation within the *All of Us* research program and are unaccounted for by our study. In addition, the use of LASSO regression in developing weights limited the size of the cohort that we could use to develop IPWps, since we could only include *All of Us* participants with complete data. Nevertheless, we were able to develop IPWps for 310,533 *All of Us* participants, which corresponds to 75% of the Controlled Tier Dataset version 7 (n=413,457). Finally, when considering the development of weights to control for volunteer participant bias, there is discussion on whether inverse probability weighting is the best method to use.^25^ In particular, participants with every low or very high probability participation will receive very high and low weights, respectively, which means that their attributes may dominate the weighted estimates.

It should also be stressed that the *All of Us* recruitment strategy was designed specifically to enhance diversity in biomedical increase the representation of groups that have been historically left out of biomedical research.^1, 4, 14^ As a result of these efforts, a number of underrepresented groups, e.g. racial and ethnic minority groups, are overrepresented in the *All of Us* cohort. Our IP weighting scheme is not intended to signal that alignment with the national population is the primary goal for *All of Us*, rather we provide IPWps as one option for researchers who may wish to increase the likelihood results of epidemiological studies using the *All of Us* cohort are externally valid with respect to the US population.

In conclusion, our study reveals the extent of volunteer participant bias in the *All of Us* cohort, while providing one potential solution in the form of IPWps that can be used to help mitigate this bias. The IPWps we develop here are made available as a community resource on the *All of Us* Researcher Workbench. Code to develop weights and analysis for this paper is available on GitHub (see Code Availability). Our subsetted cohort with the IPWps can serve to improve the representativeness of the *All of Us* cohort to study the US population. Future studies of volunteer participant bias in the *All of Us* cohort could consider other participant characteristics and/or apply different weighting schemes. As the *All of Us* cohort grows over time, and as the research community continues to work on the valuable data therein, it will be important to provide weights similar to IPWps as a way to increase confidence in the external validity of their findings.

## Data Availability

The nationally representative database used to develop weights was the 2017 – March 2020 National Health and Nutrition Examination Survey (NHANES). This data is free to access and publicly available at: https://wwwn.cdc.gov/nchs/nhanes/continuousnhanes/default.aspx?Cycle=2017-2020 The 2021-2023 NHANES sample is also free to access and publicly available at: https://wwwn.cdc.gov/nchs/nhanes/continuousnhanes/default.aspx?Cycle=2021-2023 We used version 7 of the *All of Us* Controlled Tier Dataset, which can be accessed and analyzed from the Researcher Workbench by registered users: https://www.researchallofus.org/data-tools/workbench/

## Code Availability

The code and data used to develop weights is available as a Workspace within the *All of Us* researcher workbench, which can be accessed and analyzed from the Researcher Workbench by registered users: https://www.researchallofus.org/data-tools/workbench/. The code is also available via GitHub: https://github.com/healthdisparities/Weighting-AoU.

## Data Availability

The nationally representative database used to develop weights was the 2017 - March 2020 National Health and Nutrition Examination Survey (NHANES). This data is free to access and publicly available at: https://wwwn.cdc.gov/nchs/nhanes/continuousnhanes/default.aspx?Cycle=2017-2020
The 2021-2023 NHANES sample is also free to access and publicly available at: https://wwwn.cdc.gov/nchs/nhanes/continuousnhanes/default.aspx?Cycle=2021-2023
We used version 7 of the All of Us Controlled Tier Dataset, which can be accessed and analyzed from the Researcher Workbench by registered users: https://www.researchallofus.org/data-tools/workbench/

https://wwwn.cdc.gov/nchs/nhanes/continuousnhanes/default.aspx?Cycle=2017-2020

https://www.researchallofus.org/data-tools/workbench/

## Acknowledgements

The authors would like to thank the *All of Us* volunteer participants, without whom this study and the entire project would not be possible.

We would also like to thank Dr. Christian S. Alvarez for his support in refining the imputation of the NHANES cohort and in improving the procedure for weighting the *All of Us* cohort. We would also like to thank him for critical reading of the manuscript and providing helpful feedback.

## Author Information

### Authors and Affiliations

National Institute on Minority Health and Health Disparities, National Institutes of Health, 11545 Rockville Pike, Building 11545 Rockville Pike, 2WF Room C14, Rockville, MD, 20818, USA Manoj Kambara and Leonardo Mariño-Ramírez

School of Biological Sciences, Georgia Institute of Technology, Atlanta, GA, USA Shivam Sharma and I. King Jordan

National Library of Medicine, National Institutes of Health, Bethesda, MD, USA John L. Spouge

Division of Cancer Epidemiology and Genetics, National Cancer Institute, Rockville, MD, USA Barry I. Graubard

## Contributions

IKJ and LMR conceived of the study. LMR provided supervision and funding for the *All of Us* Researcher Workbench analysis. MK carried out most of the *All of Us* Researcher Workbench data analysis and prepared the figures and tables. SS contributed to the *All of Us* Researcher Workbench data analysis. BIG and JLS supported statistical data analysis. MK and IKJ wrote and edited the manuscript. All authors read and approved of the final manuscript.

## Ethics Declarations

### Ethics approval and consent to participate

The *All of Us* operational protocol (#2016–05) is approved by the NIH Institutional Review Board. Written informed consent was obtained from all participants. All data available to researchers has had direct identifiers removed and has been further modified to minimize re-identification risks. Because the *All of Us* data were not collected specifically for this study and no one on the study team has access to the subject identifiers linked to the data, this study is not considered human subjects research according to the NIH Revised Common Rule for the Protection of Human Subjects: https://grants.nih.gov/policy/humansubjects/hs-decision.htm.

