## Supplementary information for "Increasing Representativeness in the *All of Us* Cohort Using Inverse Probability Weighting"

### **Supplementary Methods**

**Supplementary Method 1. NHANES imputation procedure**

We first ran a single multiple imputation on all variables except drinking and smoking frequency. Following the initial imputation, we ran a single multiple imputation on drinking and smoking frequency for individuals that reported drinking and smoking at least once in their lifetime. Following this we aggregated the information from both drinking and smoking questions into one variable, this aggregation is depicted in **Supplementary Table 1**. Comparison of pre- and post-imputation means and proportions for all NHANES variables indicate that the imputation of missing data was robust (R^2^ = 0.96; **Supplementary Figure 2** and **Supplementary Table 2**).

### **Supplementary Figures**

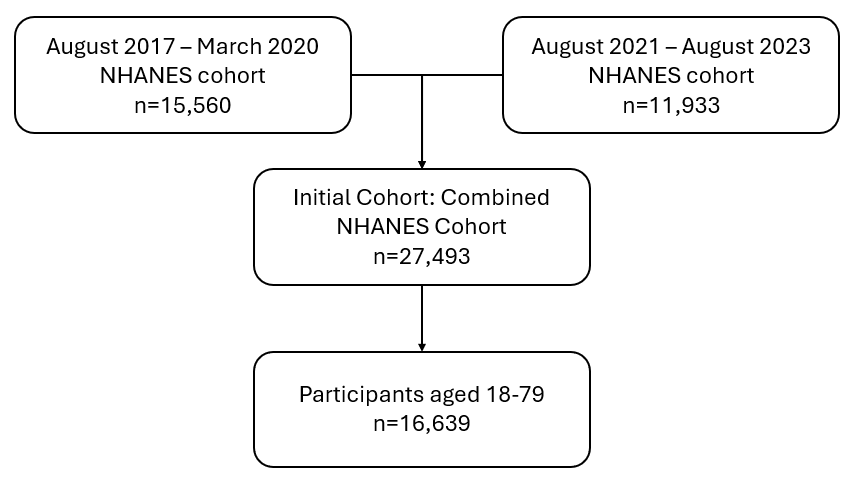

**Supplementary Figure 1. NHANES participant cohort creation.**

Inclusion criteria and the number of participants retained at each step.

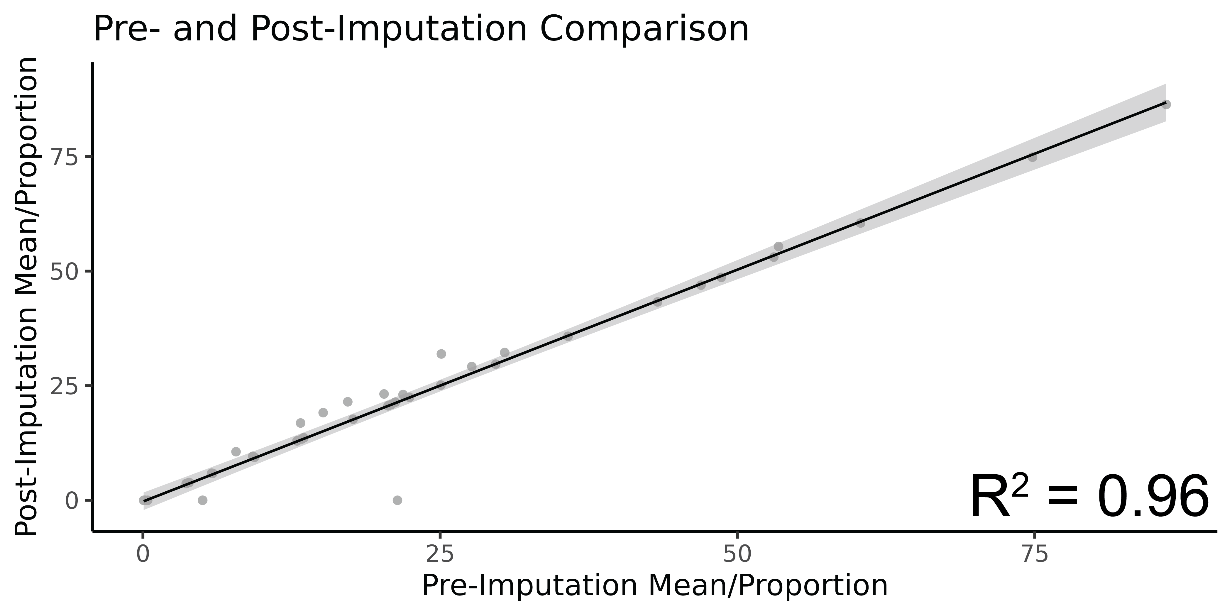

#### **Supplementary Figure 2. Pre- and post-imputation comparison.**

Graphs comparing the pre- and post-imputation means and proportions for the NHANES cohort.

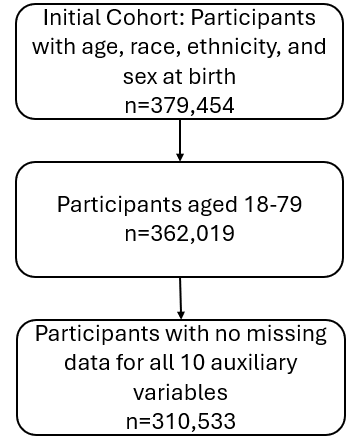

**Supplementary Figure 3. All of Us participant cohort creation.**

Inclusion criteria and the number of participants retained at each step.

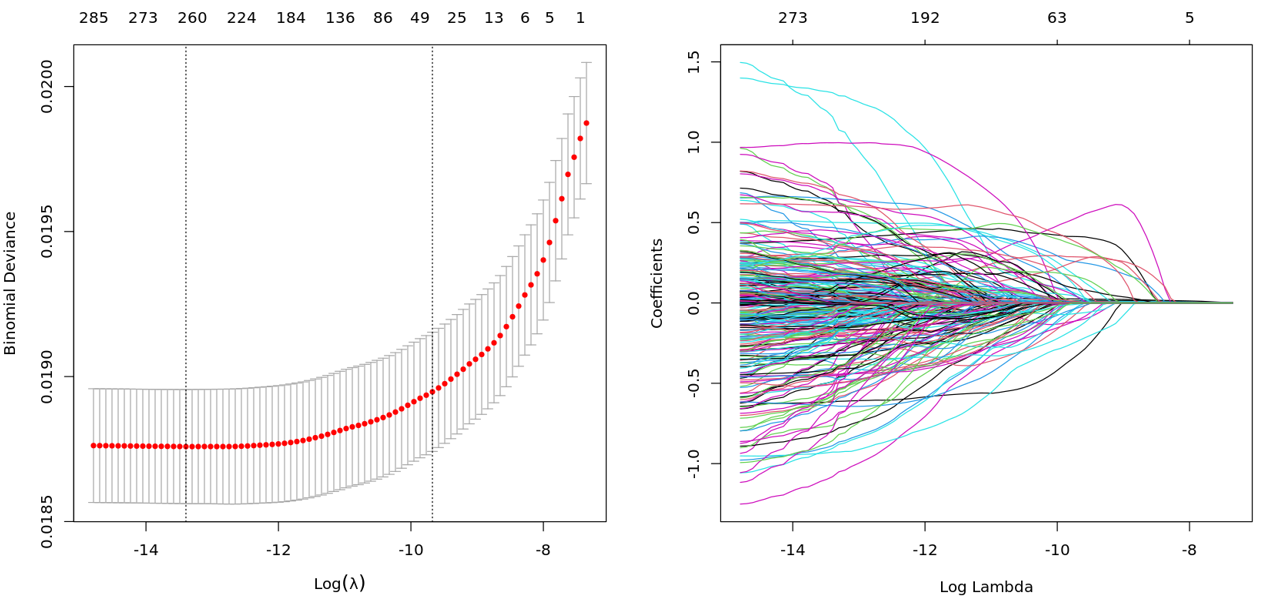

#### **Supplementary Figure 4. Graphs for LASSO regression model used for IP weight development.**

These graphs show the process of variable selection by the LASSO regression algorithm.

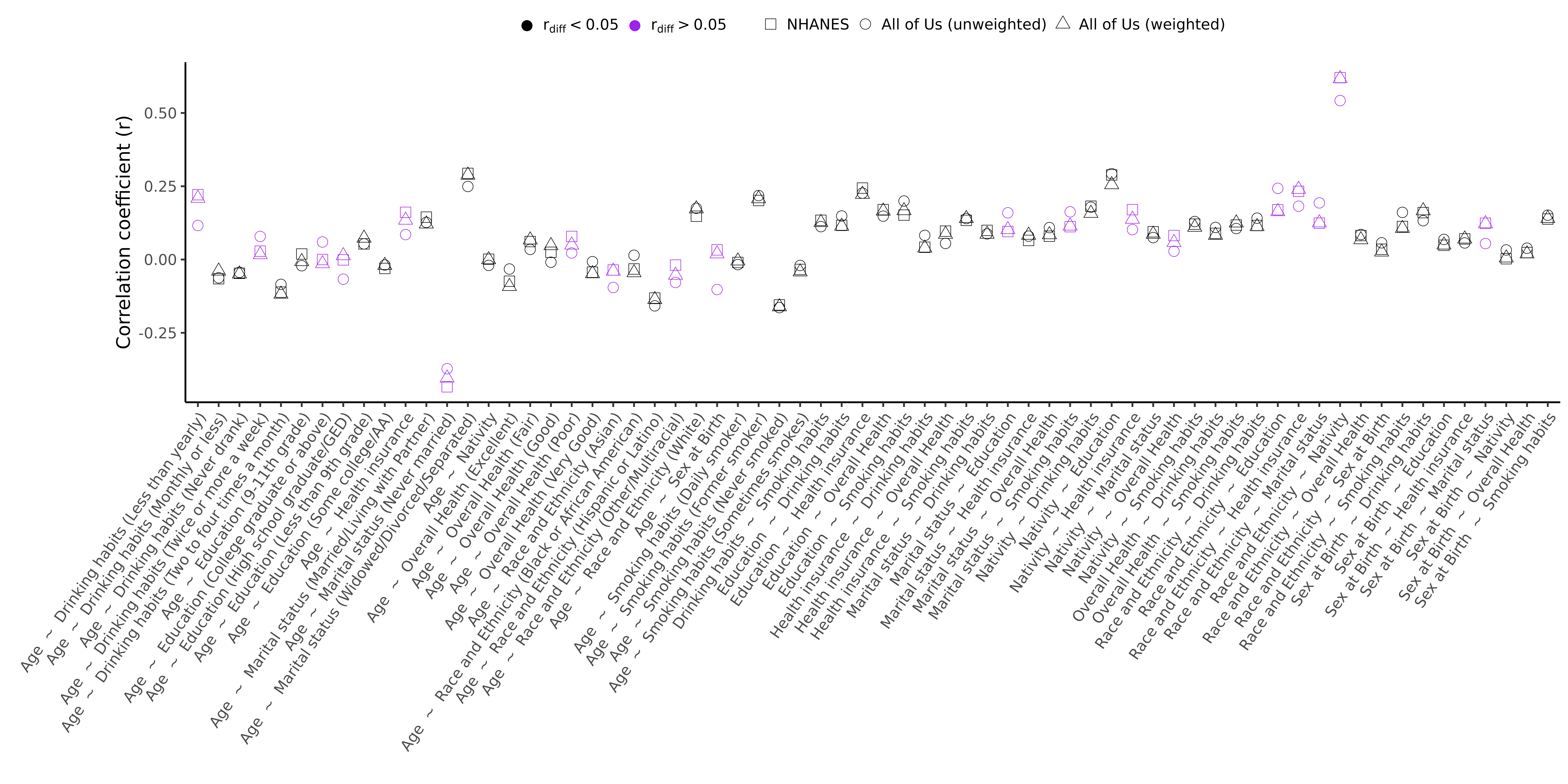

#### **Supplementary Figure 5. Correlation structure within *All of Us* data.**

The inclusion of interaction terms in the *All of Us* participation regression model allowed us to accommodate the correlation structure in the data when estimating IP weights. This can be seen when participation bias is quantified as the difference between the pairwise correlation coefficients ($r_{X.Y}$) for participant variables calculated using the NHANES or the *All of Us* cohorts [${r_{diff}=NHANES(r}_{X.Y})-AllofUs(r_{X.Y})$]. Correlation coefficients for all pairs of variables are shown for NHANES (squares), *All of Us* unweighted (circle), and *All of Us* weighted (triangle). Purple color indicates differences in correlation coefficients between NHANES and *All of Us* >0.05 prior to IP weighting.
